# Health-seeking behaviour and beliefs around sore throat in The Gambia: a qualitative study

**DOI:** 10.1101/2023.07.17.23292793

**Authors:** Maria Suau Sans, Muhammed Manneh, Isatou Ceesay, Amat Bittaye, Gabrielle de Crombrugghe, Alexander J Keeley, Thushan I. de Silva, Jennifer Palmer, Edwin P. Armitage, Michael Marks, MRCG Strep A Study Group

## Abstract

Group A *Streptococcus* (StrepA) bacteria causes a broad spectrum of diseases. The most common manifestations of StrepA infection are sore throat and pus-producing skin infections such as impetigo. Complications of StrepA infection lead to inflammation in the bones, muscles, joints, and internal organs causing acute rheumatic fever and rheumatic heart disease (RHD). In The Gambia, the RHD burden is thought to be very high. However, epidemiological data is minimal, and StrepA control programmes do not exist. This study aims to explore common beliefs and practices related to sore throats among primary caregivers of children, and healthcare providers in a community with a high StrepA disease burden. This will inform the design of preventative strategies for StrepA-related sequelae.

Four informal conversations with providers and fifteen semi-structured interviews with caregivers were conducted in the peri-urban area of Sukuta, The Gambia. Sampling was purposive and gradual, beginning from households identified to have recently experienced sore throat through a parallel cohort study. Themes explored in qualitative analysis included: sore throat causal attributions and diagnoses, care practises, health-seeking behaviour, and perceived barriers to using the biomedical sector.

Sore throats were typically perceived to affect one child in a family, disproportionately or exclusively. Sore throats are rarely perceived as life-threatening, and awareness of links between sore throat and ARF or RHD was not reported among caregivers or providers in this study population. Most cases of sore throat are initially managed at home using traditional medicine which delays resort to antibiotics, though in two instances of severe pain with the presence of exudate, fear that the child’s life was at risk prompted care-seeking through the formal health system.

Our findings can inform the development of tailored strategies to increase community knowledge of the potential long-term consequences of sore throats and appropriate care-seeking, alongside improvements in the health system, to prevent StrepA sequelae effectively.

## Background

Infection with Group A *Streptococcus* (StrepA, *S. pyogenes*) bacteria causes a broad spectrum of diseases, with the most common manifestations being sore throat and impetigo (red, pus-filled sores around the nose, mouth, hand and feet).^1,2,3^ Its transmission has been traditionally attributed to large respiratory droplets, but is yet not fully understood.^4^ StrepA infection also results in more severe complications such as acute rheumatic fever (ARF) and rheumatic heart disease (RHD), the latter being the leading cause of cardiac-related disease among people under age 25 worldwide.^5,6,7^ ARF occurs because of cross-reactivity between similar antigens found in the pathogen and the human host, primarily found in the heart and the brain.^8^ This can afflict multiple tissues and result in signs and symptoms such as polyarthritis (joint pain), erythema marginatum (pink rings on the torso and inner surfaces of the limbs), or Sydenham’s chorea (neurological disorder that causes uncontrolled movements mostly affecting hands, face and feet).^9,10^ Single or repeated episodes of ARF can cause permanent damage in the heart valves, leading to RHD.^11^

Globally, StrepA is estimated to be responsible for 18.1 million prevalent cases of severe disease.^12^ RHD alone is estimated to be responsible for 10.7 million Disability Adjusted Life-Years and 306,000 deaths annually.^13^ Sub-Saharan Africa has one of the highest RHD age-standardized prevalence and death rates worldwide, especially in urban areas.^6,14^ Yet, StrepA infections are treatable with widely available, inexpensive antibiotics, making RHD a premature and preventable cause of death.^15^ Explanations are required as to why RHD remains such a problem in Sub-Saharan Africa.

In The Gambia, many children present to hospital with RHD.^16^ However, no high-quality epidemiological data regarding the prevalence and incidence of StrepA infection are available. One study found that 53.2% of the RHD patients reported a history of recurrent sore throat but of those only 32.2% had attended a health facility, with most reporting to have used traditional medicine (TM) or nothing.^16^ In high-RHD settings such as The Gambia, many StrepA sore throats go undiagnosed and untreated. Reasons are likely to be complex involving barriers to healthcare access, perceptions of the risks and benefits of treatments and cultural understandings and norms. As children and teenagers are primarily affected by StrepA and its sequelae, a better understanding of the care seeking behaviour of caregivers of children with sore throat in such settings is therefore vital. Only two qualitative studies have been conducted on these topics (both in Uganda). They found that popular perceptions of the causes and best ways to treat sore throat varied widely from biomedical theories, and TM use was common.^17,18^ However, results from other studies cannot be extrapolated to inform tailored context-specific prevention strategies in The Gambia. To fill this research gap and to inform the design of preventative strategies for StrepA-related sequelae, we conducted a qualitative study involving informal conversations and in-depth interviews in Sukuta, The Gambia.

## Materials and methods

### Study design and procedures

We conducted a qualitative study using semi-structured in-depth interviews to explore beliefs and practices around sore throat diagnosis, care-seeking, and causes in children among Gambian caregivers and health professionals. Interviews explored understanding of sore throat causation, transmission, severity, and usual health-seeking patterns when children show signs of sore throat. The study was embedded within The Streptococcus Pyogenes Carriage Acquisition and Transmission prospective cohort study (SpyCATS - NCT05117528).

### Study setting

The study was carried out in the peri-urban area of Sukuta, in The Gambia which had a population of 47,048 and an average household size of 8.1 at the census in 2013.^19^ The main ethnic groups are Mandinka, Fula and Wolof, however, there are other minority groups. The most spoken languages are Mandinka and Wolof.

### Research team

MSS conducted the semi-structured interviews and informal conversations. Three clinical research nurses (AB, IC, and MM) with prior experience in qualitative research, assisted the primary facilitator when translations from English to Mandinka (IC, MM) or Wolof (IC, AB) were required, and vice-versa. The nurses lived and worked in Sukuta and were fluent in the local languages and English.

### Participant selection

A theoretical sampling approach was followed. Informants were purposefully and gradually selected based on emerging findings. Nurses and fieldworkers were identified as gatekeepers to the Sukuta community, knowledgeable regarding the pluralistic medical context, and informal conversations started. They were able to support identifying 13 households where children had been ‘recently’ (around six months before the study) affected by sore throats from the periodical visits they had to conduct as part of the cohort study. Informants from these interviews referred us to 2 other households they knew of not enrolled in the cohort (15 households in total), as well as a traditional healer and a drug shop vendor who were involved in treating the sore throat.

Adults of all ethnic groups and socioeconomic backgrounds were eligible. Interviewed individuals were adult (18 and over years old) caregivers, resident in Sukuta during the time of the fieldwork who had identified, experienced, or heard about an episode of sore throat in a household.

### Interviews with households

Participants were interviewed by members of the study team in English, Mandinka, Wolof or Fula as appropriate. Interviews were audio-recorded and transcribed *verbatim*. All interviews were conducted in a quiet private area, elected by the participants, and lasted between 20 and 40 minutes. The semi-structured interview guideline included sections on socio-demographics, health-seeking behaviour in response to the most recent sore throat episode, health beliefs around sore throats, triggers and barriers to care-seeking and treatment, and knowledge of ARF and RHD (S1 Appendix). The first two interviews were used to validate the guideline, which was then adjusted, and some questions reworded.

### Informal conversations and observations with providers

In addition to semi-structured interviews, non-audio recorded informal conversations were held opportunistically with three SpyCATS study nurses or fieldworkers and the pharmacy worker. The conversation with the healer was audio-recorded. Ethnographic observations were collected during routine procedures and visits to the pharmacy, traditional healer, and a herbalist stand in the Sandika herbs section of the Serrekunda Market. Notes were taken during the observations and the informal conversations, and aimed to generate hypotheses, complement the content of the interviews, and support the analysis process.

### Data management and analysis

Sociodemographic characteristics of the participants included in this study are shown in Table 1.

**Table 1.** Sociodemographic characteristics of participants at enrolment.

|  | Household interviews | Provider conversations |
| --- | --- | --- |
| **Sex** |  |  |
| Male | 3 | 4 |
| Female | 11 | 1 |
| **Age of participants** |  |  |
| 18-24 years old | 4 | - |
| 25-50 years old | 4 | 3 |
| >50 years old | 6 | 2 |
| **Occupation\*** |  |  |
| Constructor | 1 | - |
| Housewife | 3 | - |
| Cleaner | 2 | - |
| Teacher | 1 | - |
| Hairdresser | 1 | - |
| Farmer | 2 | - |
| Fieldworker | - | 1 |
| Pharmacist | - | 1 |
| Nurse | - | 2 |
| Marabout | - | 1 |
| Student | 1 | - |
| Unemployed | 2 | - |
| **Relationship with the children** |  |  |
| Mother | 7 | - |
| Father | 4 | 1 |
| Older sister | 4 | - |
| Other | - | 3 |
| **Tribal affiliation** |  |  |
| Mandinka | 9 | 3 |
| Bambara | 4 | - |
| Wolof | 1 | 1 |
| Serer | 1 | - |
| **Interview language** |  |  |
| English | 7 | 4 |
| Mandinka | 7 | - |
| Wolof | 1 | - |
| **Last level of education completed** |  |  |
| Grade 6 | 4 | - |
| Grade 8 | 3 | - |
| Grade 9 | 5 | - |
| Grade 12 | 1 | - |
| College | 1 | 4 |
| Islamic school | 1 | - |

\*One missing value

The analysis combined inductive (open coding) and deductive coding (based on themes previously identified during the interview guideline design stage and review of relevant literature) approaches. Open, axial (finding connections between codes) and selective (core category connects multiple codes) coding procedures were used.

Thematic saturation was defined as when new incoming data produced little to no new information on the topic under exploration.^20^ For some themes, saturation was reached after eight interviews, and for all but the “societal change” domain, saturation was noticed after 14 interviews. The final matrix included 37 themes grouped into three great domains.

Data from the caregivers and traditional healer was analysed separately from the biomedical care providers and presented alongside one another in the sections below, being mindful that there is not always a clear distinction between ‘biomedical’ and ‘lay’ knowledge about diseases. Illustrative quotes were included in the results section to exemplify and justify relevant themes, with viewpoints clearly labelled. To improve clarity and readability, minor grammatical changes were applied to some quotes in this report.

### Ethics and Consent

This study was approved by the joint MRC-Gambia and Ministry of Health Ethics Committee and the and the London School of Hygiene and Tropical Medicine Ethics Committee. Written informed consent was obtained from study participants (S2 Appendix). This study follows the COnsolidated criteria for REporting Qualitative research (COREQ).^21^ A completed version of the COREQ checklist is included in the supplementary material of this publication (S1 Table).

## Results

The three greater thematic domains have been used to structure the report’s findings.

### Conceptualization of sore throats and its consequences

The local terms *kankonoto dimo* (Mandinka) and *bat bumeti* (Wolof) corresponded to “*throat that is in pain*”. No local language terms were identified to refer to ARF or RHD. The words *balakando* in Mandinka or *yarambutouga* in Wolof, which correspond to fever (“hot body”), were used together with other symptom descriptions characteristic of ARF, such as joint pain, to ask participants if they had ever known anyone suffering from it or heard about the condition. The Mandinka local term *yusukusasa*, meaning cardio pathology (“heart sickness”), was used with a description of other symptoms such as joint pain to ask whether participants had heard about the RHD or known anyone who had suffered it. The English terms were also used in case they heard about the medical term.

### Case identification

Participants described sore throat as pain inside the throat that has a negative impact on eating and drinking, and sometimes swallowing saliva. Caregivers reported noticing sore throat episodes in children after they manifested their experience of pain through verbal complaints, sometimes involving crying. Some participants also described confirming the sore throat in a child upon observation of tonsillar exudates (referred to as the presence of “something white” inside the throat), or observation of lymphadenitis (“neck swallow” or “swollen glands”), which can be experienced before or simultaneously with fever (“hot body”), tiredness or otitis (“ear pain”) that initiates from the throat and “arrives*”* up to the ears:

> “I see from her she can’t swallow, she feels tired, you know, she cried like ‘I cannot swallow, my neck is paining me’” (P008, mother).

Sore throat susceptibility was not perceived to be ubiquitously distributed, with a few participants stating that “children never get it” (generally referring to babies or young children who cannot speak) or something that only certain children get. Participants from many households identified one child in the family as the sole susceptible individual, reporting single or multiple (between one and four) sore throat episodes a year:

> “Just I have only one girl who got sore throat problem, because my [other] children do not complain about sore throat. But her, only to her, sore throat, every year, two or three times” (P003, father).

If caregivers identified more than one child as being susceptible to experiencing sore throat, discussions about causality and care-seeking tended to focus on the child reporting multiple episodes.

### Perceived consequences and risks

Many participants reported being concerned about sore throat episodes in children. The most commonly emerging reason for concern was the inability to eat or drink. Unproductivity (“just lie down”), lymphadenitis, weight loss, difficulty speaking, and missing school due to sore throat, were also reported reasons for concern. Death as a perceived potential outcome of sore throat was reported twice and linked with not seeking appropriate healthcare in a health facility (“get medicines”).

> “If you don’t remove it [the exudate] so that thing would come out, it can kill someone.” (P013, older sister).

Participants who reported no concern or very mild concern classified the sore throat episodes as “normal” and easy to resolve by the usage of TM.

### Perceived causes

When it came to perceived causes of sore throat, most participants were unable to identify a causative agent for sore throat. The most widely discussed transmission model was based around the custom of members of the same compound eating with their hands from the same plate. The sore throat transmission event would specifically and consistently depend on the eaters’ hygiene-related practices after a meal-sharing event. There were variations regarding the specific hygiene-related acts identified as transmission triggers, the most common one was not washing your hands and “rubbing” them on the compound’s “wall” or “fence”.

> “When you eat [as a] family you eat from one bowl. When you wash your hand here after that you make your hand like this on the sides of the bucket, the top part. So after that, the people you [ate] with, […] our belief [is …] some will have this thing [sore throat]” (P016, mother).

> “If you eat with people in the same bowl and just wash your hand and wipe it on the fence or something then that can also cause sore throat” (P012, older sister).

Some participants also identified ingesting specific foods such as spicy chili peppers or palm oil as having the capacity to cause sore throat (in both adults and children) as well as smoking (adults only).

### Healthcare workers perceptions

The causes of sore throat, as reported by healthcare workers, were attributed to a viral or a bacterial infection. According to the pharmacist, transmission events were associated with activities such as gatherings, eating and drinking with other people and “saying lots of things” (aerosols that caused person-to-person transmission). One of the nurses said sore throat infections were caused by improper food handling (not washing or peeling the food).

In terms of identification, healthcare workers indicated that a visual inspection for “tonsil swollen” was used to differentiate between “pharyngitis or tonsilitis”, which in this context referred to irritated throat caused by a viral infection or tonsils with exudates caused by a bacterial infection. The pharmacist mentioned that if exudates were present, he would dispense Penicillin B (250mg) and Ibuprofen to customers. However, he expressed concerns about the limited resources available to conduct specific diagnostic tests, which would help avoid the unnecessary use of broad-spectrum antibiotics.

Nurses and other healthcare workers’ knowledge regarding ARF and RHD causes, identification and management were limited. When initially probed, none mentioned ARF or RHD as sequelae related to sore throats. When specifically asked about ARF and RHD, one of the nurses said ARF involved high fever and incidence followed a seasonal pattern “just like malaria”. He acknowledged lots of heart problems among children under five, with RHD being a disease that started in 2020 and that could be diagnosed by using a heart scan. Another nurse said that paediatricians and cardiologists do know about the condition, but it is “briefly explained by external speakers” during nursing studies.

### Health-seeking behaviour and care practices

A sore throat management episode would typically begin with the child experiencing it complaining to the primary caregiver (normally a mother or older sister) regarding their experience of throat pain that is hampering their food or drink intake. After the caregiver identified the situation as a sore throat episode, they would initiate actions to manage the symptoms to resolve the episode. Biomedical and traditional approaches to healthcare were often viewed as complementary.

The most frequently narrated treatment itinerary corresponded to initial use of traditional remedies at home followed by health seeking at a pharmacy or health facility if home remedies did not treat the symptoms, e.g.,:

> “we take black pepper, put it in hot water, and put it in the neck [throat]. But sometimes it doesn’t work, then we go to the hospital.” (P008, mother).

One young woman reported self-managing the condition by combining paracetamol with a *safou*, an amulet crafted by a *marabout.* Other participants indicated they never seek facility-based healthcare for a sore throat due to the perceived effectiveness of traditional remedies in solving recurrent sore throat episodes.

> “I use hot water, use it two days, it stop, all gone, so she also I did that […] only hot water in two days, it stays and three days then it stop” (P007, mother).

Within households, participants typically identified fathers as the one responsible for covering the healthcare-related expenditures, mothers and older sisters were commonly identified as chaperones to access the biomedical sector, and parents were most often identified as decision makers.

### Traditional medicine use

The most common reported home remedy consisted of mixing black pepper (*Piper nigrum)* powder in hot water, which had to be drunk or gargled up to three times a day. Licking cooking mortars that had been used to grind black pepper and ginger was also described, as was using a teaspoon covered in black pepper to either straighten the inflamed uvula (after it “collapses”) to elicit the gagging reflex in the child. The latter was used to help exudate “come out”.

For babies and young children, the *marabout* described reciting a spell and massaging the child’s neck area where “the pain is”. For older children and adults, he reported making a paste of black pepper, salt, and leaves from the mango (*Mangifera indica*) or *Gro* trees, and putting the mixture on the tongue of the patient while they inhaled steam from a bowl of hot water under a blanket. This combination was used until “the exudates” come out.

Informal conversations suggested that some people go to the market to buy herbs to treat sore throats. During a visit to a Serrekunda market herbal stand, the herbalist recommended buying a sachet containing *Banah* tree leaves powder. The *marabout* also acknowledged *Banah* had curative properties for sore throat but used mango leaves more often because they were easier to find. The pharmacist reported that such traditional medicines could have negative effects on the kidneys and the liver due to their bitterness and unknown doses being taken.

Another treatment for sore throat mentioned by a few people involved spitting over the fence or wall delimitating your compound into a compound in which you had never eaten before. Only one nurse tried to provide a causal explanation for this, suggesting that the regurgitation that predeceases the spitting act would help to remove the exudates from the tonsils.

### Biomedicine use

Following a consultation at formal healthcare facilities individuals would either receive treatment or a prescription to be taken to the pharmacy. Some participants referred to using antibiotics as superior, faster, or more appropriate for the sore throat condition. The pharmacy worker expressed concerns about antibiotic resistance and some caregiver accounts suggested they stopped giving antibiotics when symptoms resolved: “*Before those medicines finish, then the sore throat goes away. She did not take medicine again* [continue the medicine after the symptoms decreased]” (P011, mother).

### Changing societal practices

Despite the contemporaneous usage and compatibility of traditional and biomedicine, some changing patterns were reported. The most common practice change concerned the declining use of teaspoons covered in black pepper, usually executed by the elderly of the compound. This change was attributed to the discomfort elucidated by the method, compared with the simplicity of administration and perceived effectiveness of antibiotics. Another perceived cause for the subtle transition towards the preference of the biomedical sector is the death of *marabouts* with no apprentices to replace them. The practice of lymphadenectomy was also reported to be a procedure executed by *marabouts* in the past to cure and prevent future episodes of sore throats or “madness” caused by sleeping sickness which has been eliminated from this part of West Africa.

### Perceived barriers to the biomedical sector

Although the overall perception of the biomedical sector was positive, several barriers were identified.

A repeatedly identified barrier in this study were the perceived direct and indirect costs involved in seeking healthcare in the biomedical sector. Participants described having to leave their “duties” and “chores” to go to the centre, implying loss of income (indirect cost), waiting for long hours to be seen by a health worker and then being deceived when treatment was needed but not available at the same facility, meaning they had to approach a pharmacy and pay for the treatment out-of-pocket (direct cost):

> “Sometimes you leave everything, go with the sick there, from morning you’ll be in the queue until afternoon, then they will just tell you there is no medicine left, so you go and buy to the pharmacy, so then if you are there without money, how are you going to buy those medicine?” (P011, mother).

Some participants described delaying antibiotic treatment due to the need of obtaining the necessary resources to be able to buy them. The additional burden of having to go to the pharmacy after reaching the health centre was such that one participant acknowledged never using a prescription given at the health centre because of the successful resolution of her sore throat after using TM.

When participants perceived they had insufficient time to visit a health centre on the day of illness, they sometimes reported directly attending pharmacies for investigation (“checkings”) and treatment, despite increased associated costs.

Another repeatedly expressed complaint was that health care workers were perceived to have a neglectful attitude towards the patients, regardless of the severity of their conditions:

> “Sometimes these health centres, you can go there, then you see nurses there then they would refuse to attend you, they would be doing some other things. Sometimes you see a patient who is about to die and they will just be careless until the person die” (P006, father).

## Discussion

Our study provides the first assessment of beliefs about causation and care seeking behaviour for sore throat in West Africa. We identified that, in any household, sore throat is typically perceived to affect one individual, disproportionately or exclusively. Most cases of sore throat are initially managed at home using traditional medicine, with remedies consisting mainly or black pepper and water or water vapour, though healers also sometimes make a paste from local tree leaves. Only if this is ineffective is care sought through the formal health system, with most people describing receiving antibiotics. Sore throats are rarely perceived as life-threatening, and awareness of the link between sore throats and ARF or RHD was not reported in this study population.

To the best of our knowledge, this is the first in-depth analysis of a community’s understanding and health-seeking behaviour related to sore throats in The Gambia. In this study, individuals identified as suffering recurrent sore throat episodes were most commonly 5 to 14 years of age, consistent with the group described as suffering the highest burden of sore throat, ARF and RHD.^6^ Our results confirm findings from previous studies suggesting that most community members, and many healthcare workers, were unaware of the link between sore throats, ARF, and RHD and that many thought traditional remedies were appropriate to treat or manage sore throats, prior to resorting to antibiotics.^16,18,22,23^ Based on a recent review, in settings with a high risk of ARF and where point-of-care diagnostics is not available, it is recommended that all patients with acute sore throats receive antibiotics.^24^ Despite the low levels of knowledge concerning the cause and transmission of sore throats, we identified key barriers to seeking healthcare through the formal system. Both direct and indirect costs represent a significant barrier to care seeking. Whilst we did not identify evidence of catastrophic expenditure, it was clear that cost issues may delay health care seeking, increasing the possibilities of unresolved, persistent underlying infections that can lead to ARF and RHD.

Only two studies have previously conducted in-depth interviews or group discussions to understand the community’s perspective surrounding sore throat beliefs and treatment practices. Consistent with our findings, perceptions of sore throats and health-related behaviours described in those studies differed markedly from the biomedical theoretical model, with important population proportions relying upon TM use.^17,18^ In our current study some traditional remedies, such as spitting or coughing up exudates, involve actions that could actively promote the transmission of StrepA to other household members or neighbours; others such as the application of pepper and steam seem less harmful. The safety and efficacy of all of these practices could be discussed in future community engagement strategies.

Several important limitations were identified in this study. Language, gender, and cultural differences between participants and members of the research team might have affected experiences elicited due to positionality and social desirability. To address this, we worked closely with trained nurses and fieldworkers who came from the communities where the research took place and had ‘insider’ knowledge of local health discourses and traditional practices. However, working with a team of healthcare workers could have promoted social desirability bias and motivated participants to over-report biomedical sector services use. Most of the study participants were also enrolled in our SpyCATS cohort study. This is particularly relevant for the health-seeking behaviours described by the participants in the study. It is plausible that those who agreed to participate in the cohort study could contribute to a healthy participant effect, that is overall more concerned about their health status and is not necessarily representative of the whole Gambian population. The participants all lived in a single urban area in The Gambia and our findings might not represent the pluralistic culture of different Gambian regions.

## Conclusions

This is the first study to provide data on healthcare-seeking behaviours and beliefs surrounding sore throats of caregivers of children in the Gambia. Participants’ knowledge regarding causes and consequences of sore throats was low and did not necessarily prompt active healthcare seeking behaviours to the health facility nor lead a household to adopt hygiene practices that could halt asymptomatic transmission when one child tends to be sick. The findings from this study, in conjunction with the SpyCATS epidemiological outcomes, will provide a multi-disciplinary foundation for developing comprehensive interventions combining information, education, and communication strategies that, with improvements in the health system, could help prevent StrepA-related sequelae in the Gambian context.

## Data Availability

Transcripts can be reviewed by accessing the "Transcripts_collated" word document submitted for peer-reviewing purposes. This document can not be included in the final publication.

## Acknowledgements

We are grateful of the following members of the MRCG Strep A Study Group whose names are not in the main authorship list: Annette Erhart, Pierre R. Smeesters, Martin Antonio, Sona Jabang, Beate Kampmann, Anna Roca, Isatou Jagne Cox, Peggy-Estelle Tiencheu, and Grant Mackenzie. We thank the MRCG at LSHTM and study participants and the families of Sukuta for their involvement and interest in the study. We also thank Professor Koen Peteers (Institute of Tropical Medicine Antwerp) for meeting and providing MSS with theoretical resources during the study design. We also thank Franca for the generation of the first draft of the CARE form. MSS also thanks the LSHTM Trust Fund for their contribution to plane tickets to and from The Gambia.

## Authorship contribution

MSS: study design, methodology, data collection, data analysis, data interpretation, writing-original draft and editing. MMar: study conceptualization and design, supervision, resources, writing-review, editing. EPA: study conceptualization and design, supervision, coordination, writing-review and editing. JP: study design, methodology, supervision, writing-review, and editing. GC: coordination, supervision, writing-review and editing. MMan: data collection, translation, writing-review. AB: data collection, translation, writing-review. IC: data collection support, translation, writing-review. TIdS: study conceptualization, resources, writing-review and editing. AJK: writing-review and editing. All authors read and approved the final manuscript.

